# Assessing the sustainability of an integrated rural sanitation and hygiene approach: A repeated cross-sectional evaluation in 10 countries

**DOI:** 10.1101/2021.08.11.21261919

**Authors:** Paschal A. Apanga, Matthew C. Freeman, Zoe Sakas, Joshua V. Garn

## Abstract

**Introduction:** While many studies have implemented programs to increase sanitation coverage throughout the world, there are limited rigorous studies on the sustainability of these sanitation programs.

**Methods:** Between 2014 and 2018, the rural Sustainable Sanitation and Hygiene for All (SSH4A) approach was implemented by SNV in Sub-Saharan Africa and Asia. Repeated cross-sectional household surveys were administered annually throughout program implementation, and 1-to-2-years following completion of program activities. We characterize to what extent sanitation coverage was sustained 1-2 years after implementation of this SSH4A intervention.

**Results:** Surveys were conducted in 12 program areas in 10 countries, with 22,666 households receiving a post-implementation survey. Six of 12 program areas (Bhutan, Ghana, Kenya, both Nepal sites, and Tanzania) had similar coverage levels of basic sanitation 1-to-2-years post-implementation, whereas there were varied levels of slippage in the other program areas (both Ethiopia sites, Indonesia, Mozambique, Uganda, and Zambia), ranging from a drop of 63 percentage points in coverage in Ethiopia to a drop of only 4 percentage points in Indonesia. In countries that experienced losses in the coverage of household sanitation, generally sanitation sharing among neighbors did not increase, whereas open defecation did increase. In each of the areas where slippage occurred, the sanitation coverage levels at the final time point were all still higher than the initial time point before SNV started working in these areas. We found a number of factors to be associated with the sustainability of sanitation coverage, including household socio-economic status, having household members with disabilities, baseline sanitation coverage levels of the program areas, and rate of change of coverage during program activities.

**Conclusions:** Data revealed sustained gains in sanitation coverage in some program areas, yet slippage in other areas. This work may serve to benchmark sustainability of sanitation interventions in Sub-Saharan Africa and Asia.

## Introduction

The Sustainable Development Goal (SDG) target 6.2 aims to achieve access to adequate and equitable sanitation and hygiene for all and to end open defecation by 2030.^1-3^ Though it is important for sanitation programs to provide equitable access to sanitation, the sustainability (e.g., preventing stalled progress or slippage of coverage) of sanitation programs is critical^4, 5^ and has been a challenge.^6^

Slippage in sanitation coverage after completion of intervention activities has been a recognized challenge within the sector for many years.^7^ Past interventions focused on toilet access, and would typically promote one type of toilet option to all households in a village, while subsidizing a limited number of toilets for the poorest households.^8^ However, many subsidized toilets were used for other purposes or abandoned, and the approach rarely led to area-wide coverage and toilet use. Community Led Total Sanitation (CLTS)—a demand creation approach that prioritizes collective behavioral change—was developed in response to this lack of behavior change and area-wide sanitation improvements. However, CLTS also had challenges associated with sustainability, driven by the quality of latrines in some areas.^9^ Recent approaches have tried to build on the success of CLTS, while also addressing the supply side limitations. One such example is the TSSM (total sanitation and sanitation marketing) approach in Southeast Asia, that is based on CLTS and complementary social marketing techniques.^10-12^ The rural Sustainable Sanitation and Hygiene for All (SSH4A) was developed as a response to that, seeking further integration of governance and behavioral change communication components in the framework.

The SSH4A approach was developed by SNV and partners starting in 2008, in partnership with government line agencies, and subsequently tested in 5 countries in Asia.^13^ The approach aims to increase sanitation coverage by focusing on strengthening capacity around 4 components: 1) demand creation, 2) strengthening sanitation supply chain and finance, 3) hygiene behavioral change communication, and 4) water, sanitation, and hygiene (WASH) governance. Extensive country-specific programmatic adaptations and focus points of SSH4A have been published elsewhere.^14^

It is not common for sanitation projects to report on post-implementation sustainability.^15^ Many of the studies that have reported on sustainability were CLTS studies. Some of these CLTS studies found returns to open defecation 2-4 years after the completion of CLTS activities.^16, 17^ Another CLTS interventions in Ethiopia and Ghana, found open defecation increased by 8 percentage points in the year after the end of intervention activities in 1 of 4 study areas.^9^ Mukherjee and colleagues evaluated the TSSM program in Indonesia and found that communities that had achieved ODF status within 2 months of triggering TSSM activities were more likely to sustain higher coverage of improved sanitation compared to communities that took many months to achieve ODF status. ^12^ Two of the 20 communities in this TSSM program returned to open defecation between 4 and 24 months after being declared ODF. As there is a breadth of types of sanitation interventions, there is a need for studies that characterize the sustainability of broader intervention types beyond just CLTS and TSSM.

Between 2014 and 2018, the SSH4A approach was implemented in 15 countries in Sub-Saharan Africa and Asia. Evaluation of data from 11 of these countries revealed notable gains in sanitation coverage across countries.^14^ In 2018, program activities ceased and data were subsequently collected 1 or more years post-implementation. The primary purpose of this study was to assess whether the gains in sanitation coverage made by local governments/stakeholders with support of the SSH4A intervention in 10 countries were sustained 1-to-2-years after the completion of program activities. We characterize the prevalence of basic sanitation access and of various sanitation technologies over time to assess if there was slippage, sustained coverage, or gains in these variables after implementation of the intervention. We also characterized the sustainability of hygiene behaviors including prevalence of handwashing stations and child feces disposal practices. Finally, we investigated how varying community, household, and structural factors were associated with ongoing sanitation sustainability.

## Methods

### Study context

We evaluated whether attained coverage levels of key variables (basic sanitation, safe disposal of child feces, and handwashing stations) were sustained after implementation completion. The results from that previous evaluation have been published elsewhere.^14^ A post-implementation visit consisted of cross-sectional household surveys administered in 12 program areas in 10 countries at least 1year after completion of intervention activities. Study countries included Bhutan, Ethiopia (2 areas), Ghana, Indonesia, Kenya, Mozambique, Nepal (2 areas), Tanzania, Uganda, and Zambia. Program staff were no longer implementing the intervention activities but did return to administer the surveys.

### Data collection and follow-up

During this post-implementation visit, data were collected from 22,666 households between September 2019 to April 2021. The final data collection in Bhutan and Ethiopia 2 took place much later, in December 2020 and April 2021, respectively, due to delays caused by Coronavirus Disease 2019 (Covid-19) restrictions. Data from the post-implementation visits were compared to previous rounds of data collections which took place between June 2014 and January 2018, or in the case of Ethiopia 2 between May 2017 and December 2019.

The data collection process was standardized across all countries, using Akvo FLOW mobile application software.^18^ Household surveys were used to collect data on household demographic information, WASH access and use, and direct observations of WASH facilities.

As this study used repeated cross sections of randomly selected households, households within each round of data collection are not necessarily the same. Samples were drawn to be representative and comparable to previous data collections. We used the same multi-stage cluster sampling scheme that was used in the previous study to select a random and representative sample of the program areas.^14^ Due to insecurity in Uganda, 3 districts were not sampled during the third data collection round, and in order to ensure that our analyses were done in identical program areas over time, we did not include the third data collection round for Uganda here. Similarly, due to how the funding was set up in Indonesia, no data collection took place during the second round.

### Ethics

The study authors received deidentified data to perform this study. Research Integrity at the University of Nevada, Reno reviewed the study and determined that it does not require human research protection oversight by the IRB (1378687-1). SNV received approval from each of the individual countries to collect the data. Trained enumerators collected data from adult members of the household. Informed consent and protecting data anonymity were carried out by SNV for every survey.

### Outcome variables

Our primary outcome of interest was “basic sanitation,” as defined according to Joint Monitoring Program for Water Supply and Sanitation (JMP) standard classification system of sanitation technoligies.^19^ We also assessed specific sanitation technologies, including: no toilet, flush/pour flush toilet, pit latrine with slab, and pit latrine without slab/ hanging latrine. Safe disposal of child feces and access to a handwashing (HW) facility with soap were secondary outcome variables. Among households that had a child less than 3 years old, we assessed whether they safely disposed of their child feces. Access to a handwashing facility was defined as the presence of soap and water within 10 paces of a toilet.

### Predictor variables

We hypothesized that various community, household and structural factors might be associated with sanitation sustainability at the post-implementation follow-up. The predictor variables that we chose to include all had biological plausibility of potentially being associated with sanitation sustainability, based on our assessment from the literature. We used the same survey questions as had been administered in previous years of SNV’s evaluations, and also restricted to survey questions that were measured consistently between countries. We used community, household, and structural variables from the survey that were both consistently measured across countries and across time. The two community-level factors we assessed both related to the history of the previous sanitation interventions in an area. The baseline sanitation coverage of the county/district before the SSH4A intervention started was used as a predictor of post-implementation sustainability, and might be a proxy for the overall maturity of the market development, supply chains or governance systems. We also assessed if the “rate of change” in sanitation coverage from the previously implemented interventions was associated with sustainability. The “rate of change” variable compares the baseline sanitation coverage levels to the coverage levels during the final round that SNV was working in the area, and also might be related to the maturity of supply chains and commitment to sustained government involvement. Household characteristics that we included were household wealth quintile, and if there were household members with disabilities. Wealth has been associated with sanitation impact and sustainability in many studies. Wealth quintiles of each country were estimated from household assets using the EquityTool developed by the Social Franchising Metrics Working Group (https://www.equitytool.org/development/) as a guide. We compare households in the lowest two socioeconomic status (SES) quintiles to those in the upper three quintiles.^20^ We used the Washington Group short set of disability questions to construct our disability variable,^21^ which was defined as a household with a household member who reported a lot of difficulty or inability to: 1) see, 2) walk or climb steps, or 3) perform self-care such as washing or dressing. Structural factors included water table depth, soil type, tank pit location, and toilet age. Water table depth was collected using multiple depth categories, and later categorized to between 1 and 3 meters versus 3 or more meters. Higher groundwater tables (e.g., 1-3 meters) may be prone to flooding. Soil type was categorized as solid rock/clay versus other soil types. Different soil types vary in durability, and may also be used as construction materials in some countries. Toilet age was categorized as less than 1 year versus 1 year or more, and was respondent-reported.

### Analysis

To assess whether the key outcomes were sustained, we compared the program-level prevalence of outcome variables between the final round while SNV was working in the area and 1-to-2-years post-implementation; results were stratified by each of the 12 program areas. Analyses were also conducted to assess the sustainability of different technology types of the sanitation ladder, where the prevalence of various toilet types was compared between the final post-implementation survey and earlier surveys when the SSH4A approach was being implemented. We also characterize how latrine sharing changed during the 1-to-2-years following completion of program implementation.

We assessed whether community, household, and structural factors were associated with ongoing sanitation coverage by introducing interaction terms between each of these factors and post-implementation change in sanitation coverage (factor*change in sanitation coverage). Potential effect modifiers in these analyses include soil type, water table depth, toilet age, pit location, households having any person with disability, household SES, baseline sanitation coverage, rate of change in sanitation coverage from the previous four-year study. Each of these variables were assessed individually in an unadjusted model with all of the data aggregated and the only control variable was indicator variables for each of the program areas under study.

For all of the above analyses, we accounted for the stratified design and applied sampling weights to ensure representativeness with the program areas.^14^ Data were cleaned using STATA 14 SE (StataCorp, College Station, TX) and analyzed using SAS version 9.4 (SAS Institute, Cary, NC).

## Results

Six of 12 program areas (Bhutan, Ghana, Kenya, Nepal 1, Nepal 2, Tanzania) had statistically significant sustained coverage of basic sanitation at levels similar to the final round when SNV was still working in the area. There were varying levels of slippage of sanitation coverage in the other program areas (Ethiopia 1, Ethiopia 2, Indonesia, Mozambique, Uganda, Zambia), ranging from a drop of 63 percentage points in coverage in Ethiopia 1 to more modest drops (e.g., drops from 4 to 21 percentage points) in the other sites (Table 1). Among these 6 program areas that had drops in coverage, all still had higher sanitation coverage than at baseline^14^ before SNV started working in the area.

**Table 1.** Change in coverage of key variables 1-to-2-years post-implementation, shown by intervention area.

| Program areas | Basic sanitation coverage (JMP definition) |  | Safe disposal of child feces |  | HW station with soap prevalence |  |
| --- | --- | --- | --- | --- | --- | --- |
|  | Prevalence (%) in final round of SSH4A intervention (95% CI) | 1-yr % point change post-implementation (95% CI) | Prevalence (%) in final round of SSH4A intervention (95% CI) | 1-yr % point change post-implementation (95% CI) | Prevalence (%) in final round of SSH4A intervention (95% CI) | 1-yr % point change post-implementation (95% CI) |
| Bhutan | 95 (93, 97) | +2 (-1, 4) | 37 (23, 51) | -32 (-46, -18) | 64 (59, 69) | -9 (-16, -2) |
| Ethiopia 1 | 95 (95, 96) | -63 (-65, -61) | 97 (96, 99) | -60 (-64, -56) | 26 (24, 28) | -21 (-23, -19) |
| Ethiopia 2 | 99 (99, 100) | -14 (-16, -11) | 97 (95, 100) | -14 (-21, -8) | 20 (18, 23) | -10 (-13, -6) |
| Ghana | 36 (34, 38) | +3 (0, 6) | 47 (44, 50) | -5 (-9, -1) | 11 (10, 12) | -3 (-4, -1) |
| Indonesia | 95 (94, 96) | -4 (-6, -3) | 79 (74, 84) | -45 (-52, -37) | 36 (34, 39) | -4 (-7, -1) |
| Kenya | 68 (66, 69) | 0 (-3, 2) | 69 (66, 72) | -6 (-10, -2) | 10 (9, 11) | +1 (-1, 2) |
| Mozambique | 65 (62, 68) | -15 (-20, -11) | 73 (66, 80) | -8 (-18, 3) | 19 (17, 22) | +4 (0, 7) |
| Nepal 1 | 99 (98, 100) | -2 (-3, 0) | 94 (91, 98) | -16 (-26, -7) | 70 (67, 74) | +3 (-3, 8) |
| Nepal 2 | 94 (94, 95) | 0 (-1, 2) | 69 (66, 73) | +2 (-3, 7) | 76 (75, 78) | -21 (-23, -19) |
| Tanzania | 65 (63, 67) | +3 (0, 6) | 96 (94, 97) | -6 (-9, -3) | 35 (33, 37) | -27 (-29, -25) |
| Uganda | 78 (77, 79) | -21 (-24, -19) | 92 (91, 94) | +1 (-1, 3) | 4 (3, 4) | 0 (-1, 0) |
| Zambia | 91 (90, 92) | -18 (-21, -16) | 94 (92, 96) | -12 (-15, -8) | 24 (22, 26) | -21 (-23, -19) |

Many of the areas with larger drops in sanitation coverage (e.g., in Ethiopia 1, Ethiopia 2, Mozambique, Uganda and Zambia) happened where the sanitation technology being used was pit latrines with slabs. However, there was also a drop in flush/pour-flush toilets in Nepal (Figure 1). Pour flush toilets were more common in Asian countries, and less common in Sub-Saharan African countries. We did a sensitivity analysis using polytomous regression to assess the sustainability of different sanitation technologies (adjusting for SES and country) and we observed that while flush/pour-flush toilets were more likely to be sustained than other types of improved latrines, there was still slippage in the coverage of flush/pour-flush toilets between rounds 4 and 5 (model results not shown).

**Figure 1.**
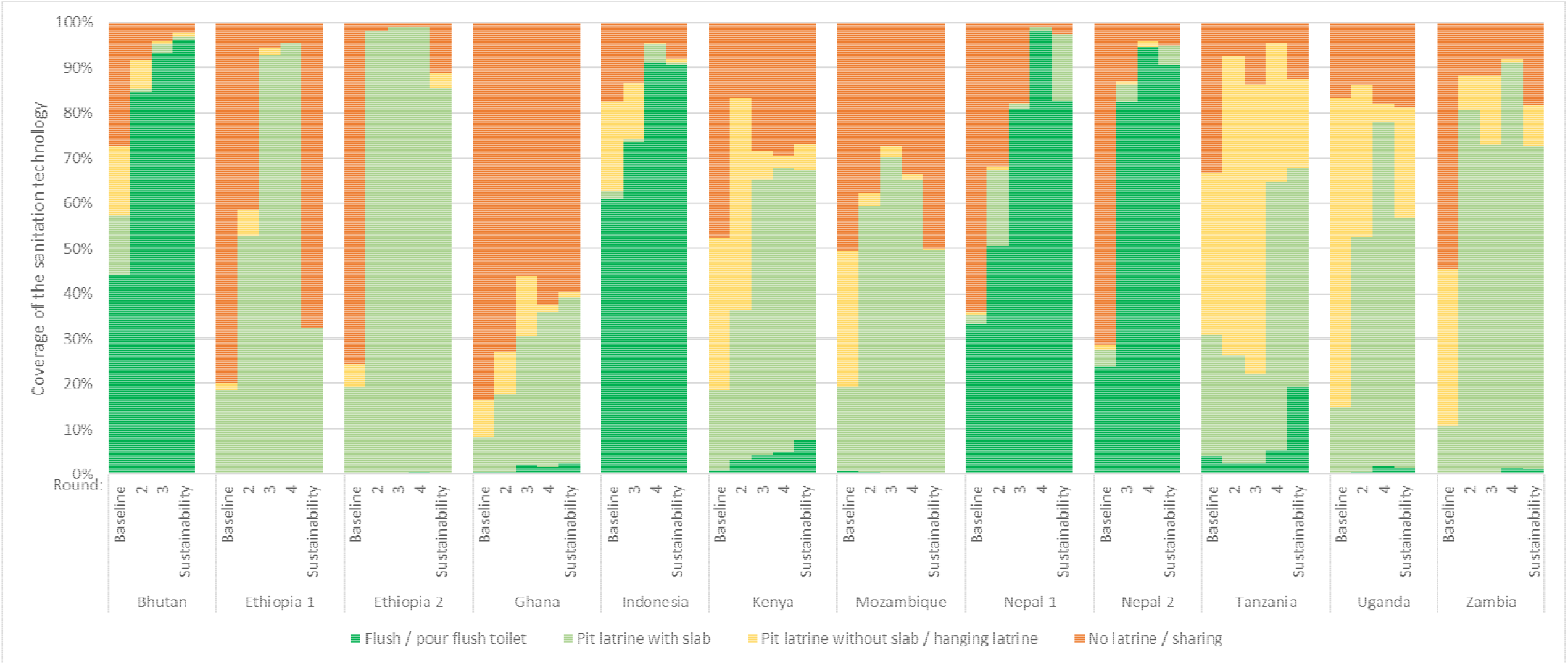
Shift in the coverage of various sanitation technology types across the 5 rounds.

Three program areas (Mozambique, Nepal 2, and Uganda) had no change in the safe disposal of child feces from the final round while SNV was working in the area to the post-implementation round (Table 1). Three program areas had less than 10 percentage points reductions in safe disposal of child feces (Ghana, Kenya, Tanzania); the other 6 program areas had drops in coverage of between 12 to 60 percentage points (Bhutan, Ethiopia 1, Ethiopia 2, Indonesia, Nepal 1, Zambia). Even though we observed that 9 program areas showed slippage in safe disposal of child feces, 8 of these areas still ended with a higher prevalence of safe disposal of child feces than they had at baseline^14^ before SNV started working in the area.

In 4 of the program areas (Kenya, Mozambique, Nepal 1 and Uganda), there was no difference in the prevalence of handwashing stations with soap at 1-to-2-years post-implementation (Table 1). The prevalence of handwashing stations with soap dropped in the other 8 program areas (Bhutan, Ethiopia 1, Ethiopia 2, Ghana, Indonesia, Nepal 2, Tanzania, Zambia) between 3 and 27 percentage points. Even though there was slippage of HW stations in 8 program areas, all these areas still ended with a higher coverage of handwashing stations with soap than they had at baseline^14^ before SNV started working in the area.

Trends in coverage of any toilet ownership, sanitation sharing, and of open defecation revealed drops in the coverage of any latrine ownership in Ethiopia 1, Ethiopia 2, Mozambique, Tanzania, and Zambia (Figure 2). In areas where there were drops in sanitation coverage, generally sanitation sharing among neighbors did not increase, whereas open defecation usually increased.

**Figure 2.**
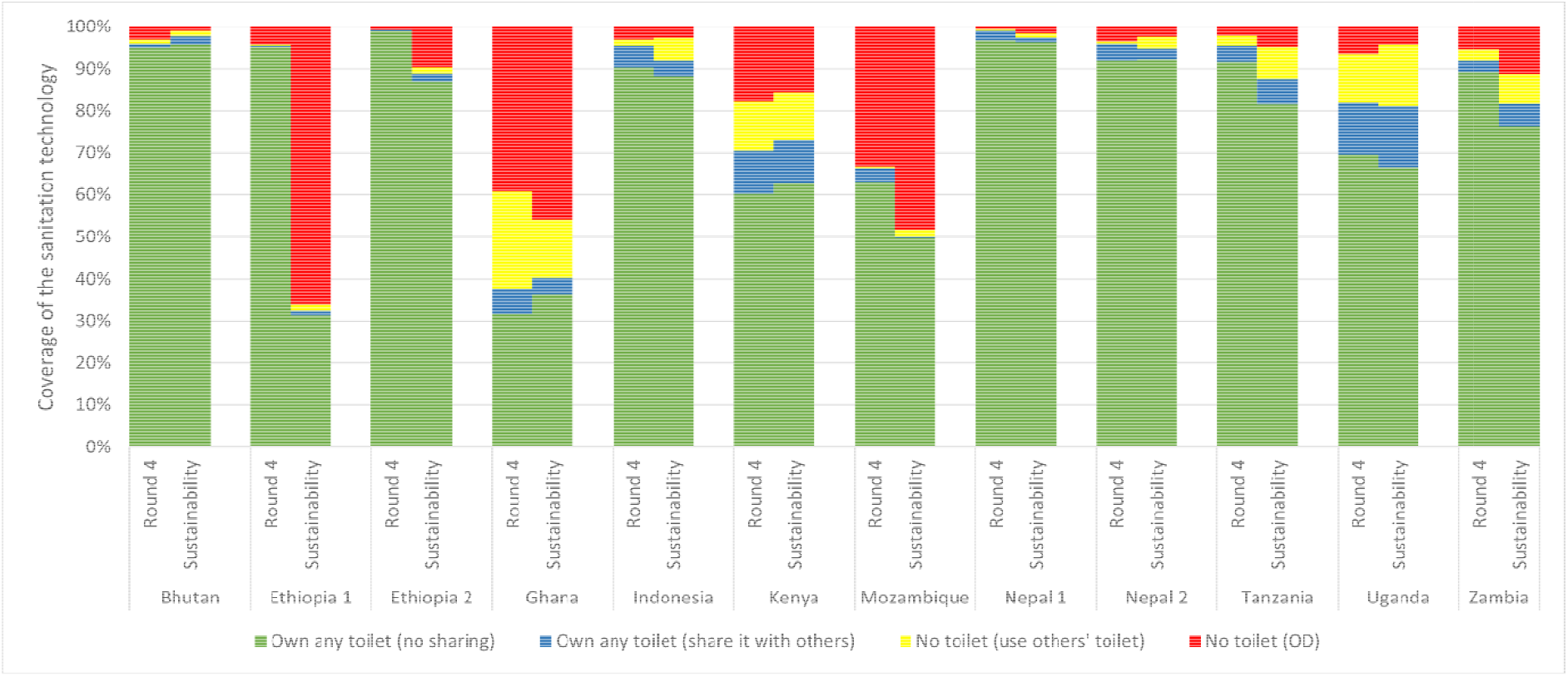
Shift in the coverage of any sanitation ownership, shared sanitation, and open defecation, comparing the final round while SNV was working in the area to 1-to-2-years post-implementation.

Data revealed several community, household, and structural factors that were associated with sustained sanitation coverage (Table 2). Households in the poorest two wealth quintiles were more likely to have had slippage of sanitation coverage (percentage point change change=-18%, 95%CI: -19%, -16%) compared to households in the upper three wealth quintiles (−11%; 95% CI: -12%, -10%). Households with persons with disabilities were less likely to have had slippage (*p=*0.03). In sensitivity analyses assessing households with persons with disabilities and slippage, but while also adjusting for potential confounders (e.g., SES and households and the household having an elderly person), the same findings persisted as in the unadjusted model (results not shown). Higher baseline sanitation coverage (i.e., higher sanitation coverage when the intervention started in 2014) was associated with better sustainability compared to having had a low baseline sanitation coverage (*p*<0.01). Having had more dramatic improvements in sanitation coverage during the previous 4-year intervention period was also associated with a higher slippage (*p*<0.01).

**Table 2.** Associations between various community, household, and structural factors and sustainability of basic sanitation (JMP definition) coverage 1-year post-implementation.

| Variable* | Post-implementation % point change in basic sanitation coverage (95% CI) | P-value |
| --- | --- | --- |
| **Household wealth quintiles** |  |  |
| Lowest two quintiles | -18 (-19, -16) | <0.01 |
| Upper three quintiles | -11 (-12, -10) |  |
| **HH with any person with disability** |  |  |
| Yes | -8 (-13, -3) | 0.03 |
| No | -14 (-15, -13) |  |
| **Baseline (round 1) sanitation coverage** |  |  |
| 75% baseline sanitation coverage | -5 (-8, -3) | <0.01 |
| 50% baseline sanitation coverage | -9 (-11, -8) |  |
| 25% baseline sanitation coverage | -13 (-14, -12) |  |
| **Rate of change in sanitation coverage from round 1 to 4 during the previous intervention** |  |  |
| +75% increase in coverage | -22 (-23, -21) | <0.01 |
| +50% increase in coverage | -11 (-12, -10) |  |
| +25% increase in coverage | 1 (-1, 2) |  |
| **Soil type** |  |  |
| Solid Rock/Clay | -13 (-14, -12) | 0.26 |
| Other | -14 (-16, -13) |  |
| **Water table depth** |  |  |
| 1-3 meter | -4 (-6, -2) | <0.01 |
| >3 meters | -15 (-16, -14) |  |
| **Tank pit above ground level\*\*** |  |  |
| Yes | -13 (-16, -11) | <0.01 |
| No | -5 (-6, -4) |  |
| **Toilet age\*\*** |  |  |
| < 1 year | -7 (-10, -5) | 0.85 |
| 1 year or more | -8 (-8, -7) |  |

\*Each variable represents a separate model that was run. Each model controlled for country program. \*\*Analysis restricted to those with a toilet. This compares the change in coverage of improved sanitation in the numerator to improved and unimproved sanitation in the denominator.

In contrast to our expectations, areas with deeper water tables were more likely to have had declines in sanitation coverage compared to households with shallower water table depths (*p*<0.01). In sensitivity analyses to explore this finding, we additionally adjusted for a number of potential confounding variables (e.g., SES), and the same findings persisted (results not shown). We also performed sensitivity analyses, stratifying to explore if this finding persisted across individual program areas, and the finding appears to be driven by results from a few select program areas (Ghana, Indonesia, Nepal 2, & Tanzania), with most program areas showing similar sustainability between deep- and high-water tables (Supplementary materials, Table S1).

There was more slippage among toilets with the tank pit above the ground level compared to toilets with tanks below the ground level (Table 2). There was no difference in slippage from improved sanitation to unimproved when comparing older toilets (≥1 year) and newer toilets (*p*=0.85; note, this analysis was restricted to participants who had a toilet).

## Discussion

We assessed the sustainability of sanitation and hygiene coverage in 12 program areas in 10 countries. The multi-country program used the SSH4A, SNV’s integrated rural sanitation and hygiene approach. Our analysis found that half of the program areas had sustained statistically similar coverage levels of basic sanitation 1-to-2-years post-implementation, whereas the other half of the program areas experienced varying degrees of slippage. Among the 6 program areas that had drops in coverage, all still had higher sanitation coverage than at baseline^14^ before SNV started working in the area. This is the first study assessing the sustainability of the SSH4A approach—an approach that has been implemented in 15 countries—and one of very few studies characterizing the sustainability of sanitation interventions.

The SSH4A approach was designed within local government planning and budgeting with the hypothesis that the improvements in sanitation and hygiene service delivery system within program areas would ensure sustainability of access and use post-implementation.^13^ Despite these efforts, some program areas did not fully sustain the intervention achievements. The finding that coverage gains were not sustained in every area, is not different from other findings in the literature, as other community-based sanitation interventions (e.g., CLTS TSSM), have reported returns to open defecation in some areas after the intervention activities ended.^12, 16, 17, 22^ Although there were also a number of program areas where sanitation coverage was fully sustained, it is not fully clear what factors might have led to high sustainability in some areas versus others.

We did find a number of community, household, and structural factors to be associated with better toilet sustainability, including households having higher socio-economic status, households having members with disabilities, living in communities that had higher baseline sanitation coverage, living in communities that had more gradual previous increases in sanitation coverage, having a tank pit below the ground, and having a toilet in an area with a shallower water table depth. Some of the community, household, and structural factors from the literature^23^ associated with sustained sanitation include seasonality,^5, 24, 25^ infrastructure,^26^ SES,^5, 16, 25-27^ education,^26^ and household structure.^28^ In contrast to our expectations, areas with deeper water tables were more likely to have had declines in sanitation coverage, but this was primarily driven by 4 program areas (Ghana, Indonesia, Nepal 2, and Tanzania), which were all sites where toilet coverage was generally sustained in our overall analyses. It is possible that during the sustainability round, each of these countries continued to build toilets, but shifted to building them in areas with higher water tables thereby changing the distribution of newer toilets to be in areas with higher water tables. Our finding on households with persons with disabilities may be explained by these households having experienced the convenience of a toilet nearby for the care of a member of disability, put a relatively higher priority to maintaining that toilet.

The literature also supports that there are several programmatic factors that can lead to increased sustainability, which include frequent personal contact with health promoters and accountability over a period of time,^23^ having an enabling environment with market access to latrine products,^26, 29, 30^ follow-up monitoring, ^26, 30-32^ social cohesion and social capital among community members,^33^ effective community leadership and political will,^31-33^ civic pride,^30^ access to sanitation markets and hardware, and sustained behavioral change.^31, 33^ One possible contributor to the high sustainability we observed in many countries may be that the SSH4A approach is an integrated, multi-dimensional approach with the aim that these different dimensions of the intervention might address the unique sanitation barriers (or enablers) in program areas with different contexts.^14^ While we did not directly assess programmatic factors associated with sustainability, some of the variables we did include, may serve as proxies for important programmatic factors. One example is baseline coverage levels. A higher coverage at baseline (before SNV arrived), might be a proxy for the overall maturity of markets, supply chains and/or governance systems in the area, which are likely to be significant facilitators/ contributors for sustained coverage in some program areas. Similarly, our findings on steep increases in coverage being associated with higher slippage, might also be related to the difficulty of sustaining coverage, sanitation supply chains and supportive governance practices in a context where overall markets and governance are weak. It may be that in some areas longer timelines and commitment may be required in order for key stakeholders to be able maintain the necessary commitment to enable sustainability.

We also observed sustained coverage for some of our secondary outcomes in some program areas. Safe disposal of child feces and access to handwashing stations with soap were sustained in 3 and 4 program areas, respectively, with varied declines in the other program areas. Notwithstanding, even in program areas that had slippage in safe disposal of child feces or handwashing stations with soap, there was almost always a higher prevalence of these variables than at baseline before SNV started working in these areas.^14^ Findings of slippage in handwashing stations, and safe disposal of child feces have previously been attributed to social norms, water shortages, climatic variability, and socio-economically disadvantaged households, and these factors have been reported as major constrains to sustained WASH behaviour.^5, 34^ Having a functional handwashing station requires a basin, water and soap, and the primary challenge with sustainability in our study was driven by the lack of soap. This was similar to finding from other studies from Ethiopia, Kenya, Uganda and Sierra Leone.^17^ Our findings of low sustainability of safe disposal of child feces in many program areas were similar to (or even better than) findings from other sustainability studies. For example, results from a CLTS program in Madagascar showed a decline in the practice of safe disposal of child feces to below baseline levels,^35^ whereas all but one of our program areas still ended with a higher prevalence of safe disposal of child feces than they had at baseline.

Our study has several limitations. The data collection periods were not always seasonally aligned, and sanitation and hygiene access may be correlated with seasonality. We also lacked data on some key contextual and programmatic variables that could be of interest to WASH sustainability (e.g., road access). As our data came from 12 program areas in 10 countries, there was considerable heterogeneity between countries, but this also may help make our results generalizable in a variety of contexts. For example, we had several program areas with low-income economies (Ethiopia, Uganda and Mozambique), one with an upper-middle income economy (Indonesia), and the remainder had low-middle-income economies. In addition, our evaluation data are from rural settings and may not be applied to all settings.

## Conclusions

The literature on sustainability from post-implementation WASH studies is limited, yet these data are critical to policymakers, program managers, and funders to characterize the success rate of programs and potentially benchmark or set targets. While our previous evaluation of the SSH4A approach found significant gains in sanitation and hygiene coverage, and in sanitation specifically,^14^ this sustainability study found that these gains did not always persist after SNV stopped working in a program area. Further rigorous studies are warranted to address how gains in coverage are, or are not, sustained. A more nuanced understanding of the sustainability of various intervention types is important for achieving universal access to adequate and equitable sanitation and hygiene, and the eventual end of open defecation.

## Data Availability

Data are available upon reasonable request.

## Role of authors

JVG and MCF conceptualized the study. JVG and PAA conducted the analysis and wrote the first draft. All authors contributed to the study design, reviewed and edited the final draft, and approved the manuscript for publication.

## Acknowledgments

This research is jointly supported by the Australian Governments Department of Foreign Affairs and Trade (DFAT), UK Department of Foreign Affairs and Trade International Development (DFATDFID) and SNV. Sustainable Sanitation and Hygiene for All is supported by: the UK Department of Foreign Affairs and International Development (DFID) in Ethiopia, Uganda, Ghana, Zambia, Kenya, Mozambique, Tanzania, Nepal; the Australian Governments Department of Foreign Affairs and Trade (DFAT) in Nepal and Bhutan; the Stone Family Foundation in Cambodia; the Embassy of the Kingdom of the Netherlands in Indonesia. We give special thanks to Antoinette Kome, Gabrielle Halcrow, Anne Mutta, and Antony Ndungu from SNV and to the data collection teams in each of the respective countries.

