## Supplementary materials, Table S1 for "Assessing the sustainability of an integrated rural sanitation and hygiene approach: A repeated cross-sectional evaluation in 10 countries"

Table S1. Unadjusted association between water table variable and sustainability of basic sanitation (JMP definition) coverage 1-to-2-years post-implementation, by country

|  | **Post-implementation % point change in basic sanitation coverage (95% CI)** | |  |
| --- | --- | --- | --- |
| *Country | Among HHs with Shallower water table | Among HHs with Deeper water table | P-value |
| Bhutan | N/A* | 2% (-1%, 4%) | N/A* |
| Ethiopia | -60% (-74%, -47%) | -63% (-65%, -61%) | 0.70 |
| Ethiopia 2 | N/A* | -14% (-17%, -12%) | N/A* |
| Ghana | 5% (2%, 8%) | -1% (-6%, 4%) | 0.03 |
| Indonesia | 1% (-3%, 5%) | -5% (-7%, -3%) | 0.01 |
| Kenya | -1% (-8%, 7%) | -1% (-4%, 1%) | 0.91 |
| Mozambique | N/A* | -15% (-20%, -11%) | N/A* |
| Nepal 1 | -2% (-5%, 0%) | -1% (-3%, 1%) | 0.46 |
| Nepal 2 | 9% (6%, 13%) | -2% (-4%, -1%) | <0.01 |
| Tanzania | 11% (5%, 17%) | 1% (-3%, 4%) | <0.01 |
| Uganda | -25% (-29%, -20%) | -20% (-23%, -18%) | 0.11 |
| Zambia | -15% (-33%, 4%) | -18% (-21%, -16%) | 0.70 |

*N/A=Not applicable, indicating that countries had a prohibitively small sample size in this stratum (Ns ranging from 0 to 5 households).
